# Correlates and Consequences of Clonal Hematopoiesis Expansion Rate: A 15-Year Longitudinal Study of 6,986 Women

**DOI:** 10.1101/2025.01.12.25320422

**Authors:** Yash Pershad, Md Mesbah Uddin, Liying Xue, Jeffrey Haessler, Jason M. Collins, Taralynn M. Mack, Elena Glick, Veronica Glaser, Kun Zhao, Siddhartha Jaiswal, JoAnn E. Manson, Urvashi Pandey, Pinkal Desai, Pradeep Natarajan, Michael C. Honigberg, Charles Kooperberg, Eric A. Whitsel, Jacob O. Kitzman, Alexander G. Bick, Alexander P. Reiner

**Author notes:** Co-supervised the work (Alexander G. Bick, Alexander P. Reiner).

## Abstract

Clonal hematopoiesis of indeterminate potential (CHIP) is associated with increased mortality and malignancy risk, yet the determinants of clonal expansion remain poorly understood. We performed sequencing at >4,000x depth of coverage for CHIP mutations in 6,986 postmenopausal women from the Women’s Health Initiative at two timepoints approximately 15 years apart. Among 3,685 mutations detected at baseline (VAF ≥ 0.5%), 50% progressed to CHIP (VAF ≥ 2%) at follow-up. We confirmed that clonal expansion is highly dependent on initial clone size and CHIP driver gene, with *SF3B1* and *JAK2* mutations exhibiting the fastest growth rate. We identified germline variants in *TERT*, *IL6R*, *TCL1A*, and *MSI2* that modulate clonal expansion rate. Measured baseline leukocyte telomere length showed differential effects on incident CHIP risk, with shorter baseline leukocyte telomere length predisposing to incident *PPM1D* mutations and longer baseline leukocyte telomere length favoring incident *DNMT3A* mutations. We discovered that the *IL6R* missense variant p.Asp358Ala specifically impairs *TET2* clonal expansion, supported by direct measurements of soluble interleukin-6 receptor and interleukin-6. Faster clonal growth rate was associated with increased risk of cytopenia, leukemia, and all-cause mortality. Notably, CHIP clonal expansion rate mediated 34.4% and 43.7% of the Clonal Hematopoiesis Risk Score’s predictive value for leukemia and all-cause mortality, respectively. These findings reveal key biological determinants of CHIP progression and suggest that incorporating growth rate measurements could enhance risk stratification.

## Introduction

Clonal hematopoiesis (CH) arises when mutations in hematopoietic stem and progenitor cells are detectable in the blood. Clonal hematopoiesis of indeterminate potential (CHIP) is defined by detection of specific preleukemic somatic mutations in peripheral blood at a variant allele fraction (VAF) > 2%.^1,2^ CHIP can be considered a pre-malignant state that occurs without overt hematologic malignancy or abnormal blood cell counts. CHIP increases risk for abnormal blood counts, subsequent malignancy, all-cause mortality, and various chronic inflammatory conditions including atherosclerotic cardiovascular disease.^3^ CHIP-associated morbidity and mortality heightens as VAF increases, so clonal expansion is a key determinant of CHIP progression to hematologic malignancy and death.^4–6^

Serial DNA sequencing is the most straightforward way to measure clonal expansion over time. However, since a single blood draw is sufficient for characterizing germline genetics, most longitudinal cohort studies perform DNA sequencing at only a single time point. A few existing longitudinal studies have produced insights into clonal behavior, primarily demonstrating variability in clonal expansion rate even among mutations sharing a driver gene.^7–12^ While studies using single blood draws have identified factors associated with prevalent CHIP, such as leukocyte telomere length (LTL), cytokine profiles, and germline variants, if and how these factors influence longitudinal clonal dynamics remains unknown.^13–15^

Using a sensitive, targeted sequencing assay, we previously characterized clonal dynamics in 548 longitudinal samples from 182 post-menopausal female participants of the Women’s Health Initiative (WHI).^9^ Here, we conducted a larger analysis of CHIP dynamics among nearly 7,000 WHI Long Life Study (LLS) participants who were sequenced at baseline and LLS examinations, roughly 15 years apart. We investigated germline genetic and environmental determinants of CHIP clonal expansion and the risks of CHIP clonal growth rate on hematological malignancy and mortality.

## Methods

### CHIP calling

The current study included 6,986 WHI LLS participants who had genomic DNA extracted at both baseline and LLS examinations (ranging from 14 to 19 years after WHI enrollment). Details of the WHI-LLS study are presented under **Supplemental Methods**. Mutations associated with CHIP were detected by single molecule molecular inversion probe sequencing (smMIPS) assay for the 11 most common CHIP genes and recurrent mutational hotspots in 4 others (**Supplemental Methods** and **Supplemental Table 1**).^9^ CHIP was defined as VAF ≥ 2%, and micro-clonal-hematopoiesis (micro-CH) was defined as VAF between 0.5% and 2%.

### Growth rate

Growth rate was estimated for each detectable mutation with VAF_base_ ≥ 0.5% at baseline and LLS using an exponential growth model: *VAF_LLS_* = *VAF_base_*(*r* + 1)*^t^* where *r* represents growth rate and *t* is the duration between baseline and LLS blood draws in years.^16^ Participants with multiple mutations were excluded, as it is not possible to reliably infer if the mutations are in the same or distinct stem cells.^13^

### Association between baseline participant characteristics, germline genetic variants, and CHIP growth rate

Baseline demographic, lifestyle, and cardiometabolic risk factors were tested for an association with clonal growth rate among those individuals with detectable CHIP at either time point (N=2,266). Association of growth rate (dependent variable) with each participant characteristic (independent variable) was assessed using linear regression adjusted for baseline age, log-transformed baseline VAF, and driver gene category as covariates.

### Germline genetic associations

Genome-wide genotyping was performed using either Affymetrix SNP 6.0 or Illumina MEGA genotyping arrays and subsequently imputed using the TOPMed reference panel (**Supplemental Methods**).^17,18^ Association with growth rate was assessed using a generalized linear model (GLM) adjusted for age at LLS, age at LLS squared, race/ethnicity, genotyping array, and the first 10 genetic principal components (PCs). The multiple hypothesis threshold for genome-wide significance was 1x10^-8^. For candidate loci previously known to be associated with CHIP prevalence by Kessler et al^19^, the threshold for significance was nominal evidence of association (P<0.05) with directionally concordant beta for CHIP growth rate and prior estimates of CHIP prevalence.

### Serum soluble interleukin-6 receptor (sIL6R) levels

Among the 2,266 women in the current study with detectable CHIP at either time point, a total of 58 had serum or plasma sIL6R levels measured in pg/mL. Z-scored serum sIL6R levels (**Supplemental Methods**) were then tested for association with growth rate for 19 women with *TET2* mutations detectable at baseline and LLS and 26 women with *DNMT3A* mutations detectable at baseline and LLS in a linear regression adjusting for age, age-squared, ancillary study, and self-identified race.

### Leukocyte telomere length (LTL) measurements

LTL was measured for a subset of 1,525 WHI participants (667 self-reported African American and 858 self-reported White) at both the baseline and LLS exams via Southern blotting (**Supplemental Methods**).^20–22^

### Cytopenia analysis

Fasting blood samples were obtained from WHI participants at baseline and LLS and were analyzed for hemoglobin, hematocrit, white blood cell count, and platelet count.^23^ We used logistic regression to examine the association between incident cytopenia (composite of anemia, thrombocytopenia, and leukopenia) between baseline and LLS and growth rate adjusting for age, race, and VAF for both baseline CHIP and baseline CHIP or micro-CH (**Supplemental Methods**).

### Time-to-event modeling

We used Cox proportional hazards regression adjusted for age at baseline, sex, smoking, and race to examine time from LLS exam to incident leukemia, lymphoma, any cancer, multiple myeloma, and all-cause mortality through February 17, 2024. In analyses of cancer, follow-up was right-censored at the time of diagnosis, death, or end of follow-up, whichever came first. Any cancer was defined as the first occurrence of any cancer (except non-melanoma skin cancer) or a death due to cancer.

### Clonal hematopoiesis risk score

The clonal hematopoiesis risk score (CHRS) at LLS exam was calculated using the criteria outlined in Table 2 of Weeks et al (**Supplemental Methods**).^6^

### Mediation analysis

We performed mediation analysis using a structural equation modeling framework to evaluate whether clonal growth rate mediated the relationship between CHRS risk at LLS and clinical outcomes (**Supplemental Methods**). CHRS categories were modeled as 0 for low-risk, 1 for intermediate-risk, and 2 for high-risk. Covariates included age, race, and baseline VAF.

## Results

### Longitudinal dynamics of clonal hematopoiesis

We performed targeted CHIP sequencing in 6,986 WHI-LLS participants using DNA obtained at baseline and LLS exams. The median ± interquartile range (IQR) of the duration between baseline and LLS sequencing was 15.8 ± 1.7 years (**Supplemental Figure 1**). Baseline characteristics of the WHI-LLS participants, stratified by presence or absence of baseline CHIP (VAF ≥ 2%), are summarized in **Supplemental Table 2**. At baseline, the median age ± IQR was 64 ± 9 years. 3,449 participants (49.4%) self-identified as non-Hispanic White, 2,160 (31.0%) as Black, and 1,105 (15.8%) as Hispanic. The prevalence of CHIP was 999/6,986 (14.3%) at baseline and increased with age to 2,275/6,986 (32.6%) at LLS. The proportion of non-*DNMT3A* mutations increased with age from baseline to LLS (**Supplemental Table 3**).

4,331/6,986 (62.0%) of participants had a mutation detected at baseline or LLS with VAF ≥ 0.5%. We examined the VAF at LLS of the 3,685 mutations detected at baseline with VAF ≥ 0.5%: 870 (23.6%) were not detected, 974 (26.4%) had 0.5% ≤ VAF_LLS_ < 2% at LLS, and 1,850 (50.2%) were CHIP (VAF_LLS_ ≥ 2%). Among baseline micro-CH (0.5% ≤ VAF ≤ 2%), 13/17 (76.5%) of *SF3B1*, 21/37 (56.8%) of *TP53*, 12/22 (54.5%) of *JAK2*, 39/74 (52.7%) of *PPM1D*, 197/431 (45.7%) of *TET2*, 23/52 (44.2%) of *ASXL1*, 503/1,645 (30.6%) of *DNMT3A*, and 42/149 (28.2%) of *ZNF318* mutations progressed to CHIP at LLS (**Figure 1A**).

**Figure 1:**
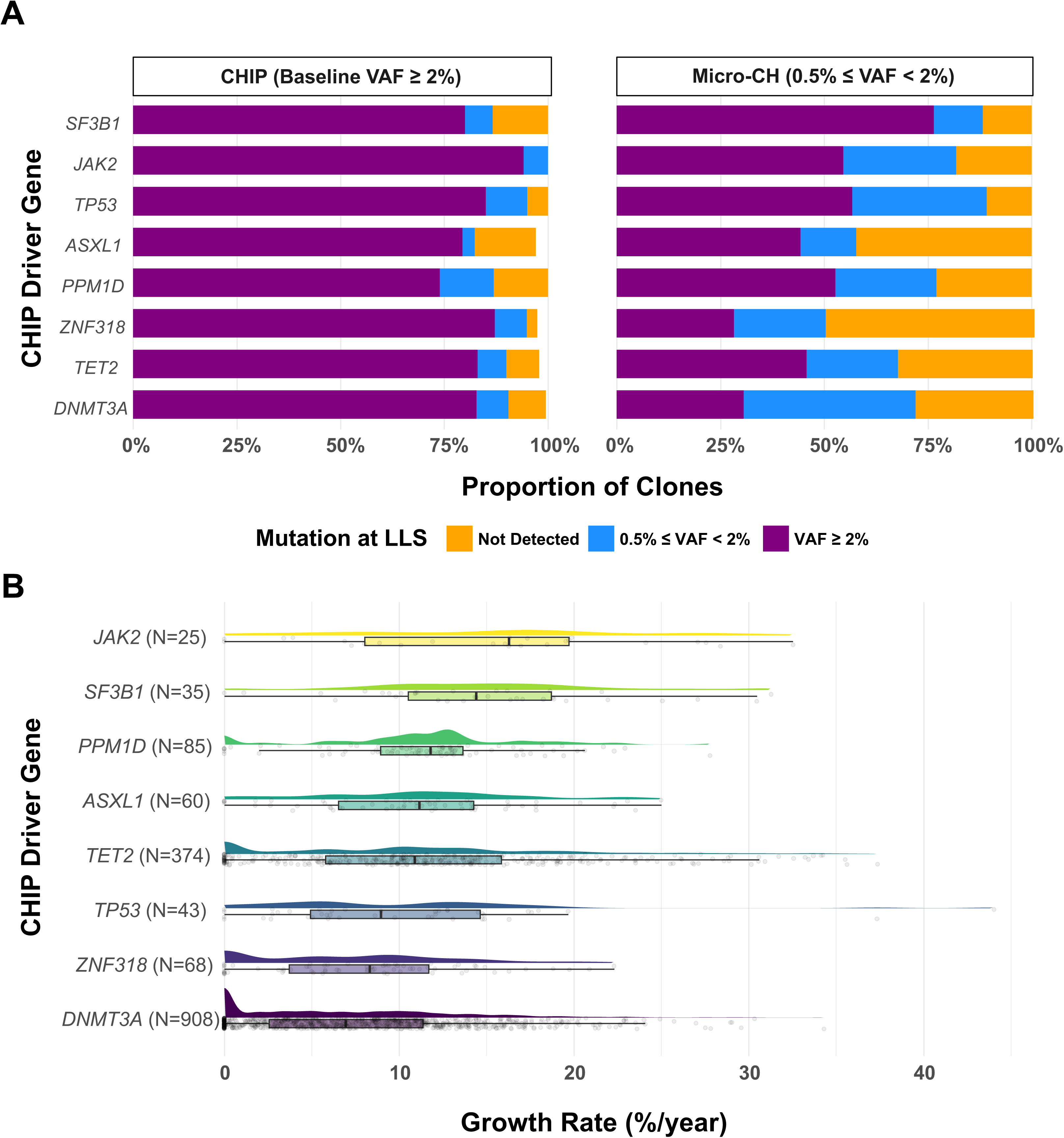
Dynamics of clonal hematopoiesis in the Women’s Health Initiative cohort. (A) Stacked barplot of mutations by driver gene stratified by VAF at follow-up sequencing. Bars show the percentage of mutations undetected (orange), with 0.5% ≤ VAF ≤ 2% (blue), or VAF ≥ 2% (purple). Left shows mutations at baseline with VAF ≥ 2%. Right shows mutations at baseline with VAF ≥ 0.5%. (B) Growth rates of CHIP mutations (VAF ≥ 2%) by driver gene. Raincloud plots show distribution, with N indicating mutations per driver gene. Mutations not detected at either time point were set to the lower limit of detection (VAF = 0.5%). Mutations with a growth rate < 0 were set as 0. Only individuals with a single mutation were included in this analysis. VAF: variant allele fraction; CHIP: clonal hematopoiesis of indeterminate potential; LLS: Women’s Health Initiative Long Life Study.

After modeling the growth rate for the 1,906 women with a single detectable mutation with VAF_base_ ≥ 0.5% at baseline and LLS (**Methods**), we found that *JAK2* and *SF3B1* mutations had the fastest annual growth rate at 16.2% and 14.4%, while *DNMT3A* and *ZNF318* had the slowest at 6.9% and 8.3% (**Figure 1B**). Micro-CH mutations (0.5% ≤ VAF < 2%) grew more slowly than CHIP (VAF ≥ 2%) mutations (median annual growth rate of 3.6% vs 9.0%, P = 4.5x10^-10^). For example, for *TET2* mutations, the median annual growth rate of CHIP was 10.9% compared to 6.0% for micro-CH (**Supplementary Table 4**).

We tested if baseline demographic and other participant characteristics were associated with CHIP growth rate. Baseline age, smoking status, race/ethnicity, education, income, region, alcohol intake, waist circumference, hip circumference, systolic and diastolic blood pressure, body mass index, creatinine, C-reactive protein, fasting glucose, insulin, HDL and LDL cholesterol, and triglycerides were not associated with growth rate after correcting for multiple hypothesis testing (0.05/number of tests) (**Supplemental Table 5**).

### Germline variants associated with CHIP prevalence modulate CHIP expansion rate

We performed two genome-wide association studies (GWASes) of growth rate among individuals with (1) CHIP and (2) CH with VAF ≥ 0.5% at either time point. No variants reached the genome-wide significance threshold in either GWAS. Since we were underpowered in our GWASes, we sought to determine the germline genetic determinants of expansion rate for candidate variants known to be associated with CHIP prevalence by a genome-wide association study from Kessler et al^19^ using a nominal significance threshold and concordance between the association direction for CHIP prevalence and growth rate (**Methods**).

We found that rs2853677, a variant in *TERT* associated with decreased CHIP prevalence,^19^ was associated with decreased CHIP clonal expansion rate (**Figure 2A**). Additional copies of the alternate allele of rs2853677 were associated with decreased CHIP growth rate (β = -0.68 %/year, 95% CI = [-0.94, -0.41], P = 8.03x10^-7^). This association was observed for *TET2* (β = -0.8 %/year, 95% CI = [-1.4, -0.2], P = 0.009), *DNMT3A* (β = -0.6 %/year, 95% CI = [-1.0, -0.2], P = 0.004), and *ASXL1* (β = -1.2 %/year, 95% CI = [-2.0, -0.4], P = 0.004). Additional copies of the alternate allele of rs2853677 were not associated with reduced growth rate of any CH mutation with VAF ≥ 0.5%.

**Figure 2:**
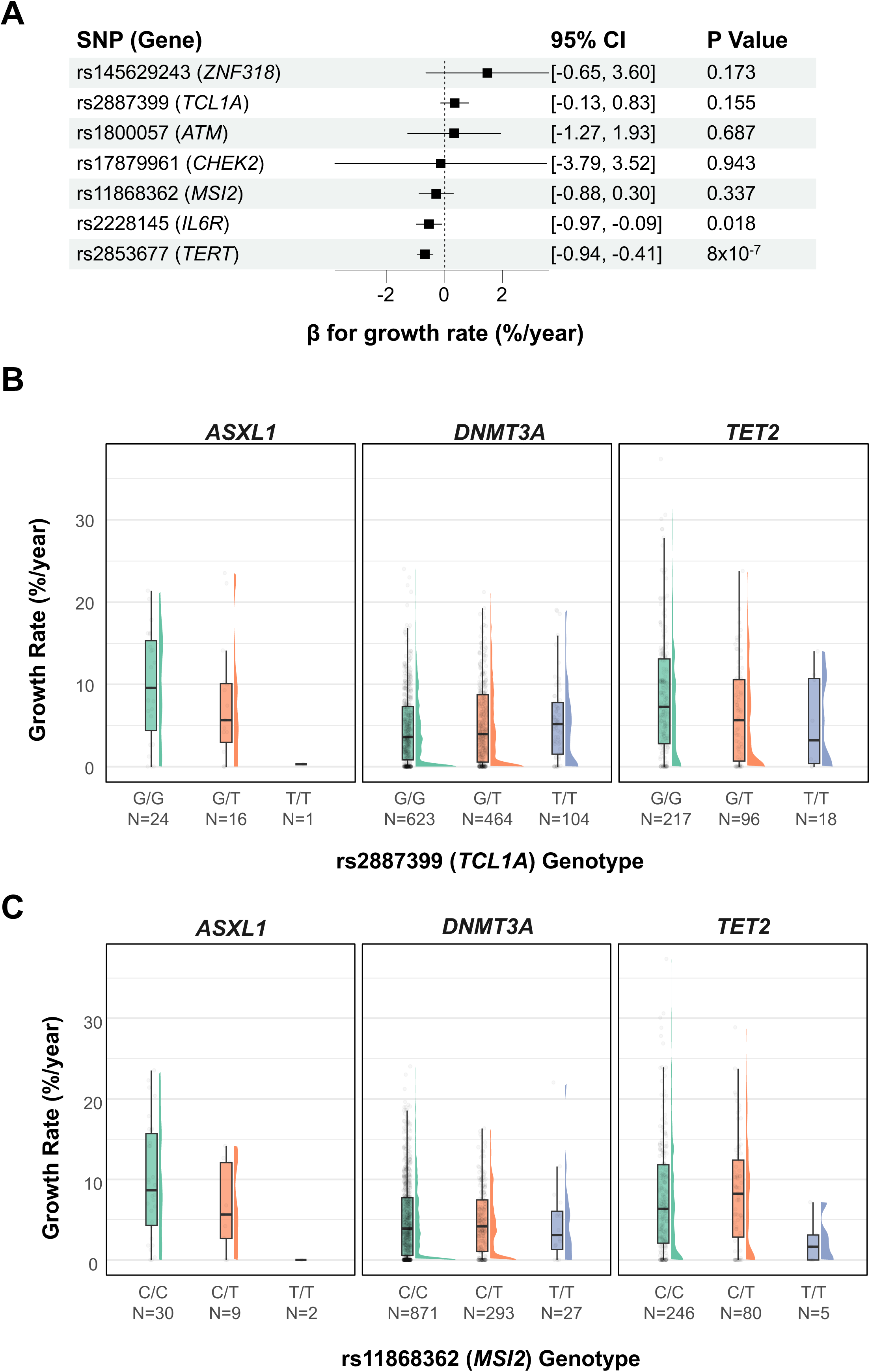
Germline genetic associations with CHIP prevalence and growth dynamics. (A) Forest plot of SNP effects on CHIP (VAF ≥ 2%) growth rate, adjusted for age, age^2^, smoking status, race, baseline VAF, and driver gene. Forest plots show effect estimates (β) with 95% confidence interval (CI) and p-values. (B) Growth rate distribution by rs2887399 (*TCL1A*) genotype for *ASXL1*, *DNMT3A*, and *TET2* mutations with VAF ≥ 0.5%. (C) Growth rate distribution by rs11868362 (*MSI2*) genotype for *ASXL1*, *DNMT3A*, and *TET2* mutations with VAF ≥ 0.5%. Forest plots show effect estimates (β) with 95% CI and p-values. In (B) and (C), the rain-cloud plot displays growth rate distributions, with sample sizes (N) for each genotype. Mutations with a growth rate < 0 were set as 0. Only individuals with a single mutation were included in this analysis. CHIP: Clonal hematopoiesis of indeterminate potential; SNP: Single nucleotide polymorphism; VAF: Variant allele fraction.

Certain genetic variants exhibited driver-gene-specific associations. Variant rs2887399 in the promoter of *TCL1A* has been previously associated with reduced *TET2* and increased *DNMT3A* clonal expansion rate, which we confirm with larger sample size of measured clonal expansion rate.^13,16^ Additional copies of the alternate allele of rs2887399 in the promoter of *TCL1A* were not associated with increased growth rate for all CHIP mutations in aggregate (β = 0.35 %/year, 95% CI = [-0.13, 0.83], P = 0.15) or any CH mutation with VAF ≥ 0.5% (β = -0.17 %/year, 95% CI = [-0.56, 0.23], P = 0.41) (**Supplemental Table 6**). However, additional copies of the alternate allele of rs2887399 were associated with increased growth rate of *DNMT3A* mutations (β = 0.73 %/year, 95% CI = [0.17, 1.2], P = 0.01, N = 826) but not for *TET2* mutations (β = 0.03 %/year, 95% CI = [-1.3, 1.3], P = 0.97, N = 341). For any CH mutation with VAF ≥ 0.5%, additional copies of the alternate allele of rs2887399 were associated with increased growth rate of *DNMT3A* mutations (β = 0.42 %/year, 95% CI = [0.24, 0.82], P = 0.04) and decreased growth rate of *TET2* mutations (β = -1.8 %/year, 95% CI = [-3.0, -0.5], P = 0.004) (**Figure 2B**).

Moreover, additional copies of the alternate allele of rs11868362 in *MSI2* – associated with reduced CHIP prevalence^19^ – were not associated with growth rate for all CHIP mutations in aggregate (β = -0.29 %/year, 95% CI = [-0.88, 0.30], P = 0.34) or any CH mutation with VAF ≥ 0.5% (β = -0.26 %/year, 95% CI = [-0.76, 0.24], P = 0.31). However, additional copies of the alternate allele of rs11868362 in *MSI2* were associated with reduced growth rate of *ASXL1* mutations with VAF ≥ 0.5% (β = -4.9 %/year, 95% CI = [-8.3, -1.4], P = 0.009) (**Figure 2C**) and suggestively associated with reduced growth rate of *ASXL1* CHIP (β = -2.5 %/year, 95% CI = [-5.3, 0.2], P = 0.07) (**Supplemental Table 6**).

### Baseline LTL associates with incident CHIP susceptibility

Given the influence of germline variants in *TERT*, which encodes the telomerase reverse transcriptase, on CHIP prevalence and growth rate, we investigated the association between measured LTL and incidence of CHIP and CHIP growth rate. First, we interrogated the effect of baseline LTL on incident CHIP. To do this, we examined the 1,293 individuals with baseline LTL measurements and without CHIP at baseline. 1,149 individuals did not have CHIP at LLS, 92 had *DNMT3A* mutations, 34 had *TET2* mutations, and 9 had *PPM1D* mutations. Shorter baseline LTL was associated with increased risk of incident *PPM1D* CHIP (OR = 1.11 per 10 kb decrease, 95% CI = [1.02, 1.21], P = 0.01), adjusted for age and race. Conversely, longer baseline LTL was associated with increased risk of incident *DNMT3A* CHIP (OR = 1.21 per 10 kb increase, 95% CI = [1.04, 1.56], P = 0.02), adjusted for age and race **(Figure 3A**). Baseline LTL was not associated with incident *TET2* CHIP.

**Figure 3:**
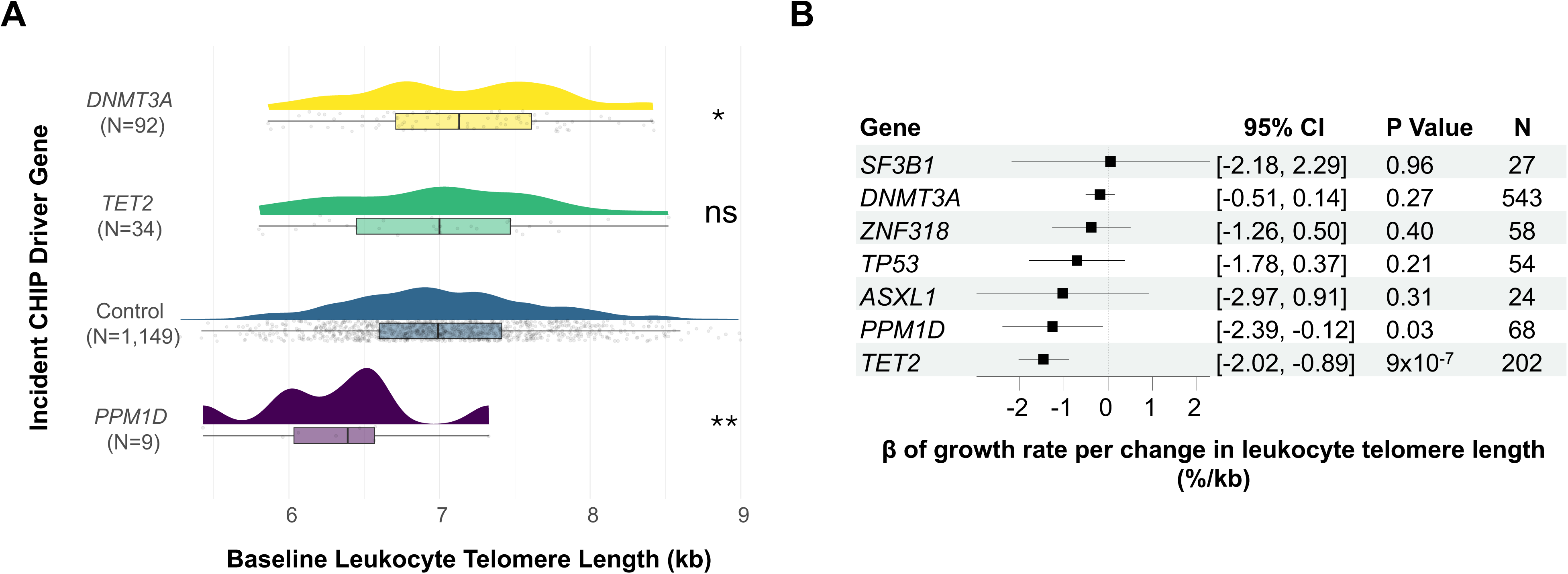
Leukocyte telomere length association with incident CHIP and growth rate. (A) Baseline leukocyte telomere length distribution by incident CHIP driver gene. Rain-cloud plots show data for individuals without CHIP at baseline (VAF < 2%). “Control” represents individuals who did not develop incident CHIP. N indicates sample size per group. *P < 0.05, **P < 0.01, ns: not significant; compared to control, adjusted for age, age², smoking status, and race. (B) Forest plot showing the association of leukocyte telomere length change growth rate for detectable CH with VAF ≥ 0.5% by driver gene. β represents the change in growth rate (%/year) per kb/year change in telomere length. 95% confidence interval (CI), p-values, and sample sizes (N) are provided. CHIP: clonal hematopoiesis of indeterminate potential; VAF: Variant allele fraction; kb: kilobase. CH: clonal hematopoiesis.

### TET2 clonal expansion associates with greater telomere shortening

Next, we examined the association between LTL and growth rate. For participants with CHIP detected at both baseline and LLS and LTL measured at baseline, baseline LTL did not associate with growth rate (P = 0.98). However, to test the hypothesis that greater telomere shortening is associated with increased CHIP growth rate, we calculated change in LTL (difference between LLS and baseline over duration between LLS and baseline in years). Among 976 participants with CHIP detected and measured LTL at baseline and LLS, faster growth rate was associated with greater LTL shortening (β = -0.90 %/kb, 95% CI = [-1.45, -0.34], P = 0.002). Faster growth rate also associated with greater LTL shortening when including micro-CH mutations; among 1,019 participants with detectable CHIP or micro-CH and measured LTL at baseline and LLS, growth rate was associated with greater LTL shortening (β = -0.74 %/kb, 95% CI = [-1.00, -0.48], P = 2.54x10^-8^). The associations were specific to *TET2* for CHIP (β = -0.93 %/kb, 95% CI = [-1.90, -0.03], P = 0.02) and any CH (β = -1.46 %/kb, 95% CI = [-2.02, -0.89], P = 9.26x10^-7^) (**Figure 3B**).

### Attenuated IL-6 signaling abrogates TET2 CHIP expansion rate

rs2228145 (p.Asp358Ala) in *IL6R* associated with reduced growth rate is a missense variant that increases sIL6R levels and reduces expression of membrane-bound IL-6R.^24,25^ This reduction in membrane-bound IL-6R impairs responses to classical IL-6 signaling on hepatocytes and leukocytes. Although not associated with CHIP prevalence in Kessler et al, rs2228145 has been shown to modify the association between CHIP and cardiovascular disease in the UK Biobank;^25,26^ we sought to test if this was due to an association with clonal expansion. We identified that additional copies of the missense variant rs2228145 (p.Asp358Ala) in *IL6R* were nominally associated with reduced CHIP growth rate (β = -0.5 %/year, 95% CI = [-0.6, -0.1], P = 0.02) (**Figure 2A**). However, additional copies of the alternate allele of rs2228145 did not associate with altered growth rate of any CH mutation with VAF ≥ 0.5% (β = -0.15 %/year, 95% CI = [-0.51, 0.22], P = 0.43) (**Supplemental Table 6**).

Measured sIL6R was available for 57 women with CHIP in WHI, of which 26 had *DNMT3A* and 19 had *TET2* CHIP. Higher sIL6R concentrations were significantly associated with decreased growth rate for *TET2* CHIP (β = -6.1 %/year per standard deviation, 95% CI = [-2.1, -10.0], P = 0.009), but not for *DNMT3A* (β = 0.46 %/year per standard deviation, 95% CI = [-3.6, 4.5], P = 0.83) (**Supplemental Figure 4**). Furthermore, higher measured IL-6 was associated with increased growth rate for *TET2* CHIP (β = 5.7 %/year per standard deviation, 95% CI = [4.2, 7.2], P = 0.01).

### Faster clonal expansion rate increases odds of incident cytopenia, leukemia, and all-cause mortality

We then sought to understand the clinical consequences of CHIP growth rate. A major clinical consequence of CHIP is abnormal blood counts – cytopenia is thought to be a critical step toward hematologic malignancy in individuals with CHIP mutations. Among individuals with CHIP at baseline and LLS, 321/2,070 (15.5%) had cytopenia at baseline and 483/2,070 (23.3%) at LLS. 333 individuals without cytopenia at baseline developed cytopenia at LLS, and 171 with cytopenia at baseline did not have cytopenia at LLS (**Figure 4A**). We next examined how growth rate in individuals with CHIP and micro-CH at baseline predicted cytopenia at LLS (**Methods**).^27,28^ Faster growth rate was associated with increased risk of cytopenia among those with CHIP (OR = 1.63 per 1%/year increase, 95% CI = [1.36, 1.95], P = 3x10^-5^) and CHIP or micro-CH (OR = 1.34 per 1%/year increase, 95% CI = [1.17, 1.53], P = 5x10^-4^), adjusted for age and race (**Figure 4B**). This increase in risk was driven by anemia and leukopenia for both CHIP and CHIP or micro-CH.

**Figure 4:**
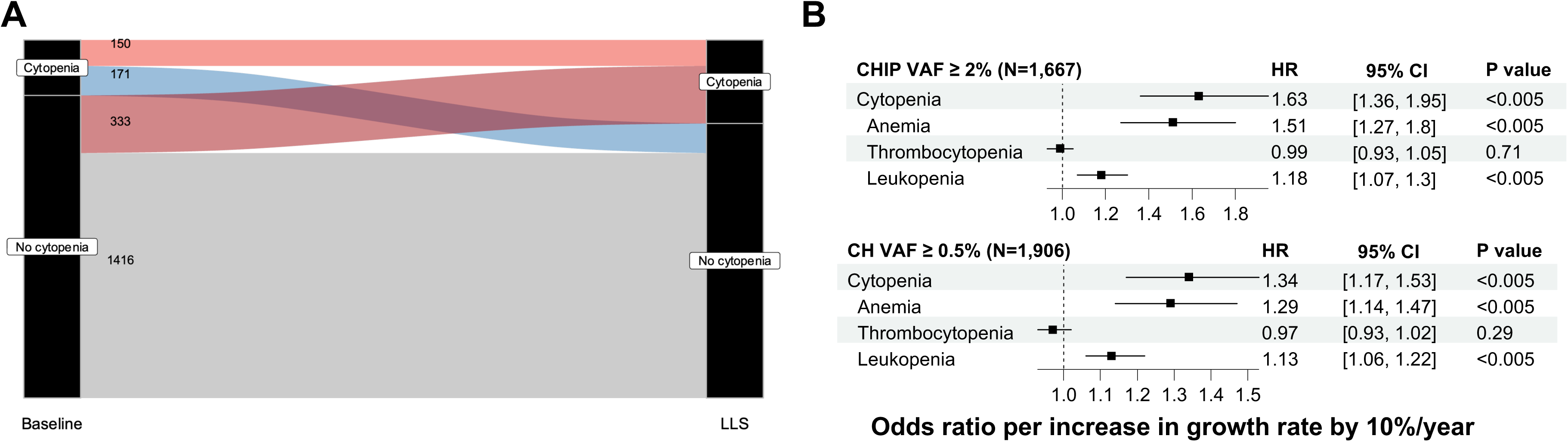
Cytopenia dynamics and CHIP growth rate associations with blood counts. (A) Sankey diagram showing cytopenia status transitions from first DNA sequencing (baseline) to second DNA sequencing (LLS). Numbers indicate individuals in each category. (B) Forest plots of CHIP growth rate effects on blood counts. Top: CHIP with VAF ≥ 2% (N=1,667); Bottom: Clonal hematopoiesis with VAF ≥ 0.5% (N=1,906). Anemia = hemoglobin concentration <12.0 g/dL (all participants were female), thrombocytopenia = platelet counts <150 × 10^9^ cells/L; and leukopenia = leukocyte count < 3.5 x 10^9^ cells/L). Cytopenia was defined as the presence of any anemia, thrombocytopenia, or leukopenia. HR represents the change in outcome risk per 1%/year increase in CHIP growth rate. 95% CI and p-values are provided. CHIP: Clonal hematopoiesis of indeterminate potential; VAF: Variant allele fraction; HR: Hazard ratio; CI: Confidence interval; LLS: Long Life Study.

We conducted time-to-event analyses to assess the association between CHIP growth rate and hematological malignancies, cancer, and mortality outcomes (**Figure 5A**). For CHIP, each 10%/year increase in growth rate was significantly associated with increased risk of leukemia (hazard ratio (HR) = 1.87, 95% CI [1.25, 2.80], P = 0.002, N = 1657) and all-cause mortality (HR = 1.09, 95% CI [1.02, 1.16], P = 0.01, N = 1665). For any detectable CH mutation with VAF ≥ 0.5%, increased growth rate by 10%/year was significantly associated with higher risk of leukemia (HR = 2.14, 95% CI [1.56-2.94], P = 2.46x10^-6^, N = 6175), lymphoma (HR = 1.71, 95% CI [1.23, 2.38], P = 1.34x10^-3^, N = 1893), and all-cause mortality (HR = 1.09, 95% CI [1.04, 1.15], P = 3.46x10^-4^, N = 1904). No significant association was found between growth rate and risk of incident multiple myeloma or any cancer (including solid organ malignancies) after correcting for multiple hypothesis testing in either VAF category. These results suggest that rate of CHIP clone expansion, particularly when considering clones of any size, is a significant predictor of hematological malignancies and mortality.

**Figure 5:**
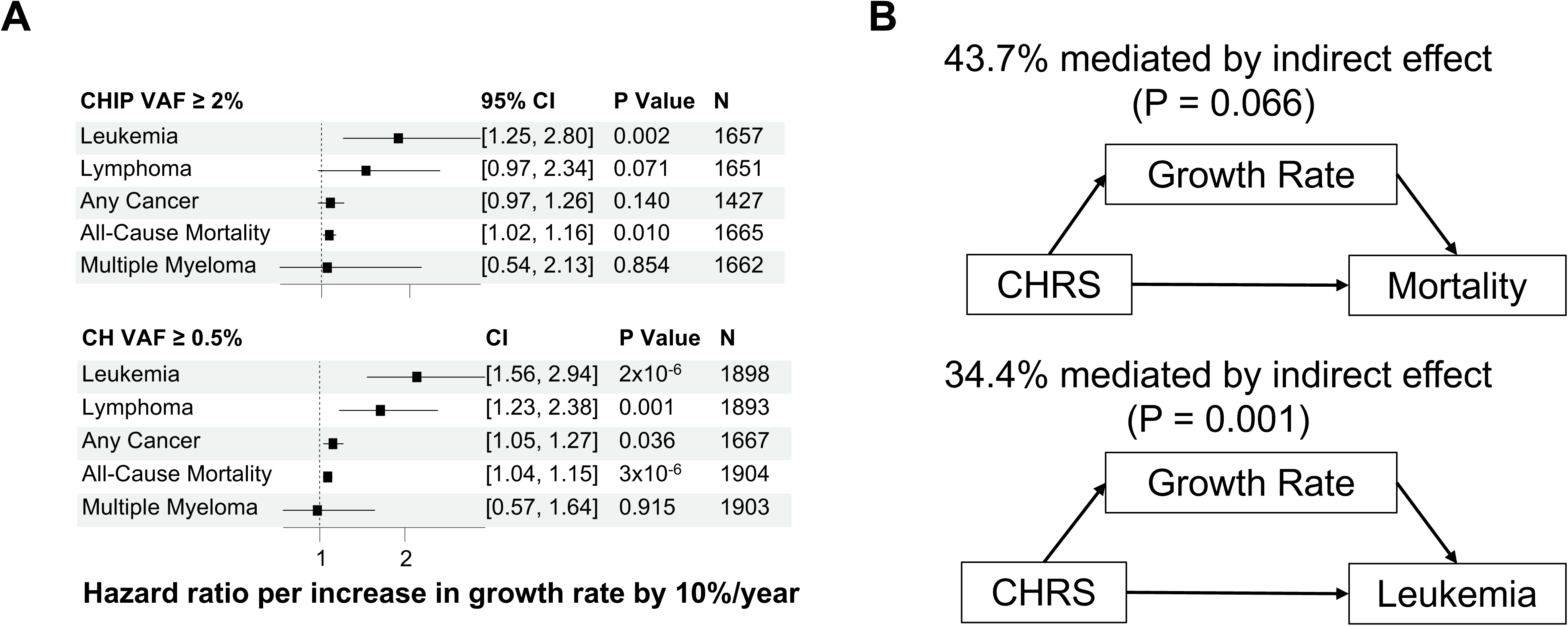
CHIP growth rate associations with hematologic malignancy and all-cause mortality and mediation with clonal hematopoiesis risk score (CHRS). (A) Forest plots showing the effect of CHIP growth rate on hema. Top: CHIP with VAF ≥ 2%; Bottom: Clonal hematopoiesis with VAF ≥ 0.5%. β represents the change in outcome risk per 1%/year increase in growth rate. 95% confidence interval (CI), p-values, and sample sizes (N) are provided. (B) Mediation analysis diagrams. Top: Growth rate mediates CHRS effect on all-cause mortality. Bottom: Growth rate mediates CHRS effect on leukemia risk. Percentages indicate the proportion of effects mediated indirectly and p-value is indicated in parentheses. CHIP: Clonal hematopoiesis of indeterminate potential; VAF: Variant allele fraction; CI: Confidence interval; CHRS: Clonal hematopoiesis risk score.

### Risk of all-cause mortality and leukemia in clonal hematopoiesis risk score is mediated by clonal expansion

We first tested whether CHRS category calculated at LLS in the WHI was associated with CHIP growth rate. After adjusting for age, race, and baseline VAF, growth rate was significantly associated with the CHRS category (β = 0.98 per 1%/year increase, 95% CI [0.80, 1.15], P = 2.0x10^-16^).

We performed mediation analysis to assess whether the association between CHRS and clinical outcomes was mediated by CHIP growth rate (**Figure 5B**; **Figure 5C**). The analysis revealed significant mediation effects for leukemia and suggestive effects for all-cause mortality. For leukemia, 34.4% of the total effect of CHRS was mediated by growth rate (indirect effect β = 0.019, P = 0.001; direct effect β = 0.019, P < 0.001; total effect β = 0.054, P = 0.018; N = 1657). For all-cause mortality, 43.7% of the total effect was mediated by growth rate (indirect effect β = 0.040, P = 0.066; direct effect β = 0.041, P = 0.062; total effect β = 0.092, P = 0.306; N = 1665). The mediation effect for any cancer was notably weaker and non-significant (20.4% mediated, indirect effect P = 0.591, N = 1427). These findings suggest that CHIP growth rate partially mediates the relationship between CHRS and subsequent risk of leukemia, with a similar trend observed for all-cause mortality.

## Discussion

Here, we used a sensitive assay to detect CHIP mutations at two timepoints with a lower limit detection of VAF of 0.5% in 6,986 women from the Women’s Health Initiative. Then, we characterized genetic and biological determinants of clonal expansion rate in CHIP and their clinical consequences. We identified germline variants in *TERT, IL6R*, *TCL1A*, and *MSI2* which modulate both growth rate of CHIP and micro-CH, the role of IL-6 signaling in *TET2* clonal expansion, and demonstrated that clonal dynamics partially mediate the prognostic value of clonal hematopoiesis on risk of incident leukemia and mortality. Our analyses enable four key insights about longitudinal dynamics of clonal hematopoiesis.

First, clonal hematopoiesis mutations are ubiquitous but many mutations do not progress to CHIP (VAF ≥ 2%). While clonal hematopoiesis mutations were detected in 62% of women, among clonal hematopoiesis mutations detected at baseline with a VAF ≥ 0.5%, about 20% were not detectable 15 years later, around 25% remained below the 2% VAF threshold for CHIP, and approximately 50% progressed to CHIP. We cannot profile these mutations in larger biobanks, as clones below VAF of 2% are not detectable by 30x depth-of-coverage whole genome sequencing.^4,29^ Moreover, these data provide important context for risk stratification: although both *TP53* and *SF3B1* mutations are considered “high-risk”, a small *SF3B1* clone has a higher likelihood of transforming to CHIP than a small *TP53* clone (75% versus 55%). A smaller *DNMT3A* clone has a 30% chance of progressing to CHIP. Prior work on longitudinal CHIP with targeted sequencing has demonstrated similar growth rates and progression from smaller clones to CHIP.^7,12,16^

Second, our findings establish telomere biology as a key determinant of CHIP prevalence and expansion. Mendelian randomization has demonstrated that individuals with genetically longer telomeres have an increased predisposition to clonal hematopoiesis of indeterminate potential (CHIP), which in turn accelerates telomere shortening.^14^ We identified a *TERT* variant which affects CHIP prevalence and growth rate, suggesting that telomerase activity influences not only CHIP mutation acquisition, but also clonal expansion. Interestingly, the effect of the *TERT* variant is much stronger for CHIP than any CH ≥ 0.5%, in contrast to the *TCL1A* and *MSI2* variants. This discrepancy for the *TERT* variant between CHIP and any CH ≥ 0.5% – despite additional sample size when including mutations with lower VAF – may suggest that telomere length only becomes constraining once the clone has reached an appreciable size (i.e., after a large number of replication cycles).

Further, this germline genetic evidence is supported by measured LTL. We observed that baseline LTL differentially affected the emergence of specific CHIP drivers – shorter LTL predisposed to incident *PPM1D* mutations, while longer LTL favored incident *DNMT3A* mutations. These data are in concordance with evidence that those with pathologically short telomeres from telomere biology disorder are predisposed to *PPM1D* CHIP.^30^ Further, we show that LTL not only predisposes individuals to develop CHIP, but also changes with clonal expansion. *TET2* clonal expansion rate was associated with accelerated telomere shortening. To-date, no work has investigated the relationship between CHIP clonal expansion and measured LTL dynamics on a per-driver gene basis.

Third, we uncovered a novel role for IL-6 signaling in regulating *TET2* CHIP expansion rate through complementary genetic and molecular evidence. The *IL6R* missense variant p.Asp358Ala, which attenuates classical IL-6 signaling by increasing sIL6R levels analogously to tocilizumab 4-8 mg/kg infusions every 4 weeks^24^, was associated with reduced clonal growth despite not being previously associated with *TET2* CHIP prevalence. This genetic association was corroborated by direct measurements showing higher sIL6R levels were associated specifically with reduced *TET2* mutation expansion while higher IL-6 levels were associated with greater *TET2* mutation expansion. IL-6 signaling has been implicated by prior work in *TET2* CHIP. Shin et al demonstrated that treatment of macaques with a weekly dose of 10 mg/kg tocilizumab for 4 months reduced VAF of *TET2* CHIP mutations.^31^ Furthermore, *IL6R* p.Asp358Ala has been shown to protect individuals with *TET2* CHIP from atherosclerosis.^25,26^ These data suggest that the protective effect of the missense variant on cardiovascular disease in *TET2* CHIP may be due to reduced clonal expansion rate. Moreover, our genetic evidence – in conjunction with prior biologic plausibility – suggests that drugs targeted IL-6 such as tocilizumab or olamkicept may be an effective way to slow *TET2* clonal expansion rate.

Fourth, our analysis reveals that clonal growth rate captures much of the prognostic information currently derived from static CHIP measurements in risk prediction models. The clonal hematopoiesis risk score (CHRS) incorporates multiple features including age, blood counts, and mutation characteristics to predict risk of myeloid neoplasm and mortality.^6^ Our mediation analysis demonstrated that growth rate accounts for 34.4% and 43.7% of CHRS’s predictive value for leukemia and mortality respectively. This analysis suggests that many established risk factors - including age-related changes in blood counts and mutation burden - may ultimately reflect differences in clonal expansion rate. The strong relationship between growth rate and clinical outcomes supports incorporating longitudinal clone dynamics into risk assessment when available.

These findings should be interpreted in the context of several limitations. Our study population consisted exclusively of postmenopausal women, predominantly of White and Black ancestry, potentially limiting generalizability to men and other populations. While our 15-year follow-up provides valuable insights into clonal dynamics, we only had two timepoints for all participants. Our targeted sequencing approach, while cost-effective for large-scale longitudinal analysis, may have missed mutations in genes not included in our panel. Our sample size for certain analyses, particularly those involving measured sIL6R levels (n=57) and LTL (n=1,525), was limited compared to our overall cohort. Moreover, certain measurements were only available in subsets of the WHI LLS participants. Women with sIL6R levels measured were either fracture, multiple myeloma, and colorectal cancer cases or their matched controls due to the design of the WHI ancillary studies in which sIL6R was originally measured. Similarly, women with LTL measurements were cases of incident cardiovascular disease and death and their matched controls.^21^ Finally, the lack of large, deeply phenotyped longitudinal studies of CHIP clonal growth with LTL measurements and cytokine measurements limits our ability to replicate our findings, but demonstrates the novelty of this cohort study.

The insights derived from this work have important implications for both understanding CHIP biology and improving clinical management of CHIP. The identification of specific pathways regulating clonal expansion, particularly IL-6 signaling and telomere maintenance, suggests potential therapeutic targets. Moreover, our findings indicate that measuring or estimating clonal growth rate could enhance risk stratification beyond current methods. Future studies should examine whether interventions targeting these pathways can modify CHIP progression and its associated complications.

## Data Availability

All data produced in the present study are available online via the WHI dbGaP accession number (phs000200.v12.p3).

## Acknowledgments

This work was supported by NIH grants DP5 OD029586 and R01 AG088657, a Burroughs Wellcome Fund Career Award for Medical Scientists, an E.P. Evans Foundation grant, a RUNX1 Research Program grant, a Pew Charitable Trusts and Alexander and Margaret Stewart Trust Pew-Stewart Scholar for Cancer Research award (A.G.B.); NIH grant T32 GM007347 (Y.P.); and NIH grants R01 HL148565 (A.P.R., E.A.W). The WHI program is funded by the National Heart, Lung, and Blood Institute, National Institutes of Health, U.S. Department of Health and Human Services through contracts 75N92021D00001, 75N92021D00002, 75N92021D00003, 75N92021D00004, 75N92021D00005.

## Author Contributions

Y.P., A.P.R., A.G.B., J.K., E.A.W., C.K., P.N., S.J., J.E.M., U.P., P.D., and M.C.H. conceived of the study. T.M.M. and K.Z. helped design statistical analysis. J.K., M.M.U., and L.X. performed CHIP detection. Y.P. and A.P.R. performed statistical analyses and wrote the manuscript. J.H. performed the genome-wide association study. All authors reviewed and revised the manuscript. A.G.B and A.P.R. jointly supervised the work.

## Disclosures and Conflicts of Interest

A.G.B. has received honoraria for advisory board membership from, and holds equity in, TenSixteen Bio. P.N. reports research grants from Allelica, Amgen, Apple, Boston Scientific, Genentech / Roche, and Novartis, personal fees from Allelica, Apple, AstraZeneca, Blackstone Life Sciences, Bristol Myers Squibb, Creative Education Concepts, CRISPR Therapeutics, Eli Lilly & Co, Esperion Therapeutics, Foresite Capital, Foresite Labs, Genentech / Roche, GV, HeartFlow, Magnet Biomedicine, Merck, Novartis, Novo Nordisk, TenSixteen Bio, and Tourmaline Bio, equity in Bolt, Candela, Mercury, MyOme, Parameter Health, Preciseli, and TenSixteen Bio, and spousal employment at Vertex Pharmaceuticals, all unrelated to the present work.

## Notes

### Author Declarations

Human samples were obtained with informed consent. Fred Hutchinson Cancer Center Institutional Review Board (IRB #10186) reviewed our study and gave ethical approval for this work

## References

1. Steensma, D. P. et al. Clonal hematopoiesis of indeterminate potential and its distinction from myelodysplastic syndromes. Blood 126, 9–16 (2015).

2. Li, W. The 5th Edition of the World Health Organization Classification of Hematolymphoid Tumors. In Leukemia (ed. Li, W.) (Exon Publications, Brisbane (AU), 2022).

3. Jaiswal, S. et al. Age-Related Clonal Hematopoiesis Associated with Adverse Outcomes. N Engl J Med 371, 2488–2498 (2014).

4. Vlasschaert, C., et al. A practical approach to curate clonal hematopoiesis of indeterminate potential in human genetic datasets. Blood blood.2022018825 (2023) doi:10.1182/blood.2022018825.

5. Vlasschaert, C., et al. CHIP Is Associated with Cardiovascular Disease in the UK Biobank. http://medrxiv.org/lookup/doi/10.1101/2023.11.30.23299001 (2023) doi:10.1101/2023.11.30.23299001.

6. Weeks, L. D., et al. Prediction of Risk for Myeloid Malignancy in Clonal Hematopoiesis. NEJM Evidence 2, (2023).

7. Fabre, M. A. et al. The longitudinal dynamics and natural history of clonal haematopoiesis. Nature 606, 335–342 (2022).

8. Mack, T. et al. Cost-Effective and Scalable Clonal Hematopoiesis Assay Provides Insight into Clonal Dynamics. The Journal of Molecular Diagnostics 26, 563–573 (2024).

9. Uddin, M. M. et al. Longitudinal profiling of clonal hematopoiesis provides insight into clonal dynamics. Immun Ageing 19, 23 (2022).

10. Van Zeventer, I. A. et al. Evolutionary landscape of clonal hematopoiesis in 3,359 individuals from the general population. Cancer Cell 41, 1017–1031.e4 (2023).

11. Robertson, N. A. et al. Longitudinal dynamics of clonal hematopoiesis identifies gene-specific fitness effects. Nat Med 28, 1439–1446 (2022).

12. Uddin, M. M. et al. Long-term longitudinal analysis of 4,187 participants reveals insights into determinants of clonal hematopoiesis. Nat Commun 15, 7858 (2024).

13. Weinstock, J. S. et al. Aberrant activation of TCL1A promotes stem cell expansion in clonal haematopoiesis. Nature 616, 755–763 (2023).

14. Nakao, T. et al. Mendelian randomization supports bidirectional causality between telomere length and clonal hematopoiesis of indeterminate potential. Sci. Adv. 8, eabl6579 (2022).

15. Mack, T. M. et al. Epigenetic and proteomic signatures associate with clonal hematopoiesis expansion rate. Nat Aging 4, 1043–1052 (2024).

16. Mack, T. et al. Germline genetics, disease, and exposure to medication influence longitudinal dynamics of clonal hematopoiesis. haematol (2024) doi:10.3324/haematol.2024.286513.

17. Bien, S. A. et al. Strategies for Enriching Variant Coverage in Candidate Disease Loci on a Multiethnic Genotyping Array. PLoS One 11, e0167758 (2016).

18. Taliun, D. et al. Sequencing of 53,831 diverse genomes from the NHLBI TOPMed Program. Nature 590, 290–299 (2021).

19. Kessler, M. D. et al. Common and rare variant associations with clonal haematopoiesis phenotypes. Nature 612, 301–309 (2022).

20. Shadyab, A. H. et al. Associations of Accelerometer-Measured and Self-Reported Sedentary Time With Leukocyte Telomere Length in Older Women. Am. J. Epidemiol. amjepid;kww196v1 (2017) doi:10.1093/aje/kww196.

21. Carty, C. L. et al. Leukocyte Telomere Length and Risks of Incident Coronary Heart Disease and Mortality in a Racially Diverse Population of Postmenopausal Women. ATVB 35, 2225–2231 (2015).

22. Shadyab, A. H. et al. Association of Accelerometer-Measured Physical Activity With Leukocyte Telomere Length Among Older Women. The Journals of Gerontology: Series A 72, 1532–1537 (2017).

23. Eaton, C. B. et al. The cross-sectional relationship of hemoglobin levels and functional outcomes in women with self-reported osteoarthritis: results from the Women’s Health Initiative. Semin Arthritis Rheum 41, 406–414 (2011).

24. Ferreira, R. C. et al. Functional IL6R 358Ala allele impairs classical IL-6 receptor signaling and influences risk of diverse inflammatory diseases. PLoS Genet 9, e1003444 (2013).

25. Bick, A. G. et al. Genetic Interleukin 6 Signaling Deficiency Attenuates Cardiovascular Risk in Clonal Hematopoiesis. Circulation 141, 124–131 (2020).

26. Vlasschaert, C., Heimlich, J. B., Rauh, M. J., Natarajan, P. & Bick, A. G. Interleukin-6 Receptor Polymorphism Attenuates Clonal Hematopoiesis-Mediated Coronary Artery Disease Risk Among 451 180 Individuals in the UK Biobank. Circulation 147, 358–360 (2023).

27. Brogan, J. et al. Risk of Incident Cytopenia in Clonal Hematopoiesis. Preprint at 10.1101/2024.09.30.24314665 (2024).

28. Cargo, C. et al. Predicting cytopenias, progression, and survival in patients with clonal cytopenia of undetermined significance: a prospective cohort study. The Lancet Haematology 11, e51–e61 (2024).

29. Bick, A. G. et al. Inherited causes of clonal haematopoiesis in 97,691 whole genomes. Nature 586, 763–768 (2020).

30. Gutierrez-Rodrigues, F. et al. Clonal landscape and clinical outcomes of telomere biology disorders: somatic rescue and cancer mutations. Blood 144, 2402–2416 (2024).

31. Shin, T.-H. et al. A macaque clonal hematopoiesis model demonstrates expansion of TET2-disrupted clones and utility for testing interventions. Blood 140, 1774–1789 (2022).

