## Supplemental Methods and Figures for "Correlates and Consequences of Clonal Hematopoiesis Expansion Rate: A 15-Year Longitudinal Study of 6,986 Women"

### *Women’s Health Initiative (WHI) and WHI Long Life Study (LLS)*

The WHI is a long-term, prospective study focused on strategies for preventing chronic diseases in postmenopausal women. The WHI study design, eligibility criteria, recruitment methods, and measurement protocols have been described previously. Briefly, the WHI collected data on 161,808 post-menopausal women aged 50–79 years (including 17% from under-represented minority populations) who were enrolled between 1993 and 1998 at 40 geographically diverse clinical centers throughout the United States. The WHI study received institutional review board approval with informed consent from all participating clinical centers. WHI participants were enrolled in either the observational study (n = 93,676) or randomized to at least one of three overlapping clinical trials (hormone therapy, dietary modification, and calcium/vitamin D supplements; n=68,132). At enrollment, WHI participants completed self-administered questionnaires covering demographics; general health; clinical and anthropometric characteristics; functional status; healthcare behaviors; reproductive, medical, and family history; personal habits; medication use; and dietary intake. WHI has accumulated large numbers of clinical outcomes via interview and event surveillance following standardized protocols. Incident cancers and deaths were reviewed, classified, and adjudicated during WHI follow-up by physician adjudicators using medical records including hospital discharge summaries, ICD-9 codes, diagnostic, laboratory, surgical, and pathology reports.

The main analyses for the current study were performed in a subset of WHI participants in the WHI LLS conducted during 2012–2013 (ranging from 14 to 19 years after WHI enrollment). The LLS involved an in-person visit, phlebotomy, measurement of anthropometrics, blood pressure, and physical function. The LLS included former participants of the WHI hormone trial, as well as African American and Hispanic women. Women who resided in an institution or were unable to provide informed consent due to dementia were excluded from the LLS. Of 14,081 WHI participants who were eligible for the LLS, 7,875 women, 63–99 years of age, successfully completed the LLS at-home visit.

*smMIPs assay and CHIP detection*

A smMIPS capture panel tiled coding exons (+/- 5 base pairs) of the 11 most common CHIP genes and recurrent mutational hotspots in 4 others (**Supplemental Table 1**). Of the 6,986 LLS participants, 13,790/13,972 samples (98.7%) were sequenced successfully at both time points (median depth of coverage, 4,609x).

Somatic variants were called from smMIPS sequencing using VarScan2, and variants were annotated using ANNOVAR software. CH were called using a Workflow Description Language pipeline (<https://app.terra.bio/#workspaces/terra-outreach/CHIP-Detection-Mutect2>). Variants were filtered if total coverage (DP) <100, supporting reads for alternative allele (AD) <10, forward and reverse reads <2, and variant allele fraction is <0.001. Scripts used for CH calling and filtering are available in <https://github.com/MMesbahU/clonalhematopoiesis-in-whi>.

*Germline genetic associations*

SNPs were filtered for minor allele frequency ≥1%, call rate ≥ 95%, HWE p-value > 10^-6^, as well as exclusion of sites with invalid or mismatched alleles for the reference panel. The data were imputed separately using the TOPMed imputation server and combined to run the GWAS. The analyses were run in R using GLM. Results were reported for SNPs with MAF ≥ 0.001.

### *Serum soluble interleukin-6 receptor levels*

A subset of WHI participants (total N=4,865 of the original 161,808 enrolled) had serum or plasma soluble interleukin-6 receptor (sIL6-R) levels in pg/mL measured by ELISA or Luminex assay [Quantikine HS Human Immunoassay DR600 (R&D Systems); Milliplex Human Cytokine / Chemokine Panel (Luminex Xmap] as part of three ancillary studies: biochemical antecedents of fracture in minority women (BA9), IGF and multiple myeloma (207), and pro- and anti-inflammatory cytokines and colorectal cancer (208). Among the subset of 2,266 women in the current study with detectable CHIP (VAF ≥ 0.02) at either time point, a total of 58 had soluble IL-6 receptor levels measured. Serum soluble interleukin-6 receptor levels were normalized and converted to z-scores for all participants of each ancillary study and then combined across ancillary studies. These z-scores were then tested for correlation with growth rate for 19 women with *TET2* mutations detectable at baseline and LLS and 26 women with *DNMT3A* mutations detectable at baseline and LLS in a linear regression adjusting for age, age-squared, ancillary study, and self-identified race.

*Leukocyte telomere length measurements*

Leukocyte telomere length (LTL) was measured in the WHI. LTL in kilobases (kb) was measured in duplicate at both time points by the mean length terminal restriction fragments using Southern blotting. Mean LTL was used for analysis for baseline and LLS. Individuals with LTL values exceeding 3 standard deviations from the sample mean were excluded from analyses.

*Cytopenia analysis*

At the LLS examination, a full CBC was performed. As per World Health Organization criteria, anemia was defined as hemoglobin < 12 g/dL, thrombocytopenia as platelet count < 150x10^9^ cells/L, and leukopenia as leukocyte count < 3.7x10^9^ cells/L. Individuals were defined as cytopenic at LLS if they had anemia, thrombocytopenia, or leukopenia at LLS. We quantified the percentage of individuals with cytopenia at LLS among those with detectable mutations at baseline.

*Clonal hematopoiesis risk score*

Prior work by Weeks et al has described the Clonal Hematopoiesis Risk Score (CHRS), which stratifies risk of CHIP transformation into leukemia and all-cause mortality in the UK Biobank. The CHRS uses features such as mutation count, driver gene, VAF, age, cytopenia, and cell morphology to prognosticate risk, but it does not incorporate CHIP longitudinal dynamics. Participants were called low-risk for CHRS ≤ 9.5, intermediate-risk for 10 ≤ CHRS ≤ 12, and high-risk CHRS ≥ 12.5.

*Mediation Analysis*

We performed mediation analysis using the lavaan (v.0.6-19) structural equation modeling framework in R. For leukemia and all-cause mortality, we fitted a two-equation model: a mediator equation (CHRS category ~ growth rate + covariates) and an outcome equation (event ~ CHRS risk + growth rate + covariates + log(time)). The model estimated *direct (c), indirect (ab), and total (c + ab) e*ffects, where 'a' represents the effect of growth rate on CHRS risk, and 'b' represents the effect of CHRS risk on the outcome. Time was incorporated as a log-transformed offset term in the outcome equation. Only observations where the blood draw preceded the event or censoring were included in the analysis.

**Supplemental Figures**

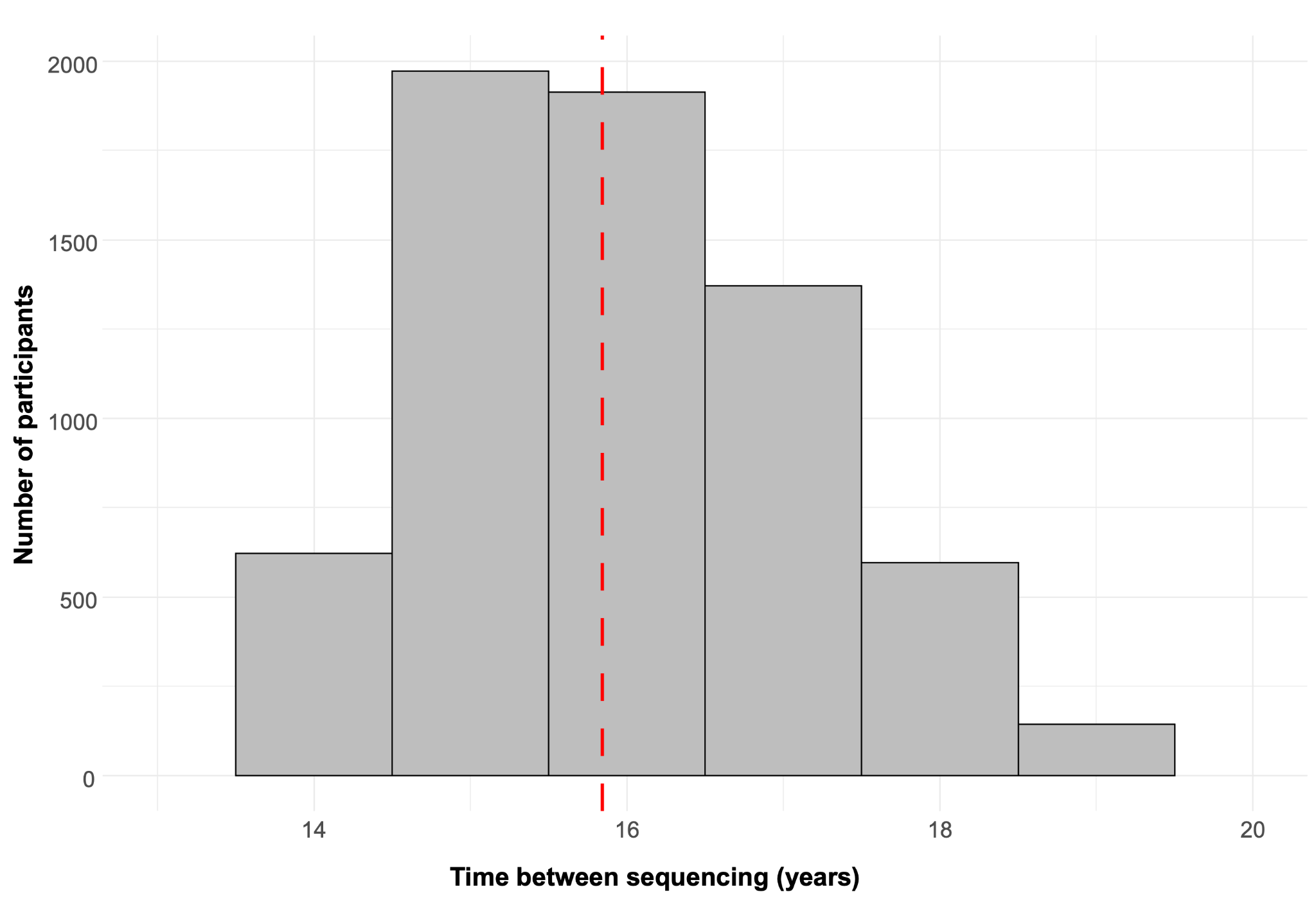

**Supplementary Figure 1: Duration between baseline sequencing and sequencing for the Long Life Study (LLS).** Median of 15.8 years is marked with a red line.

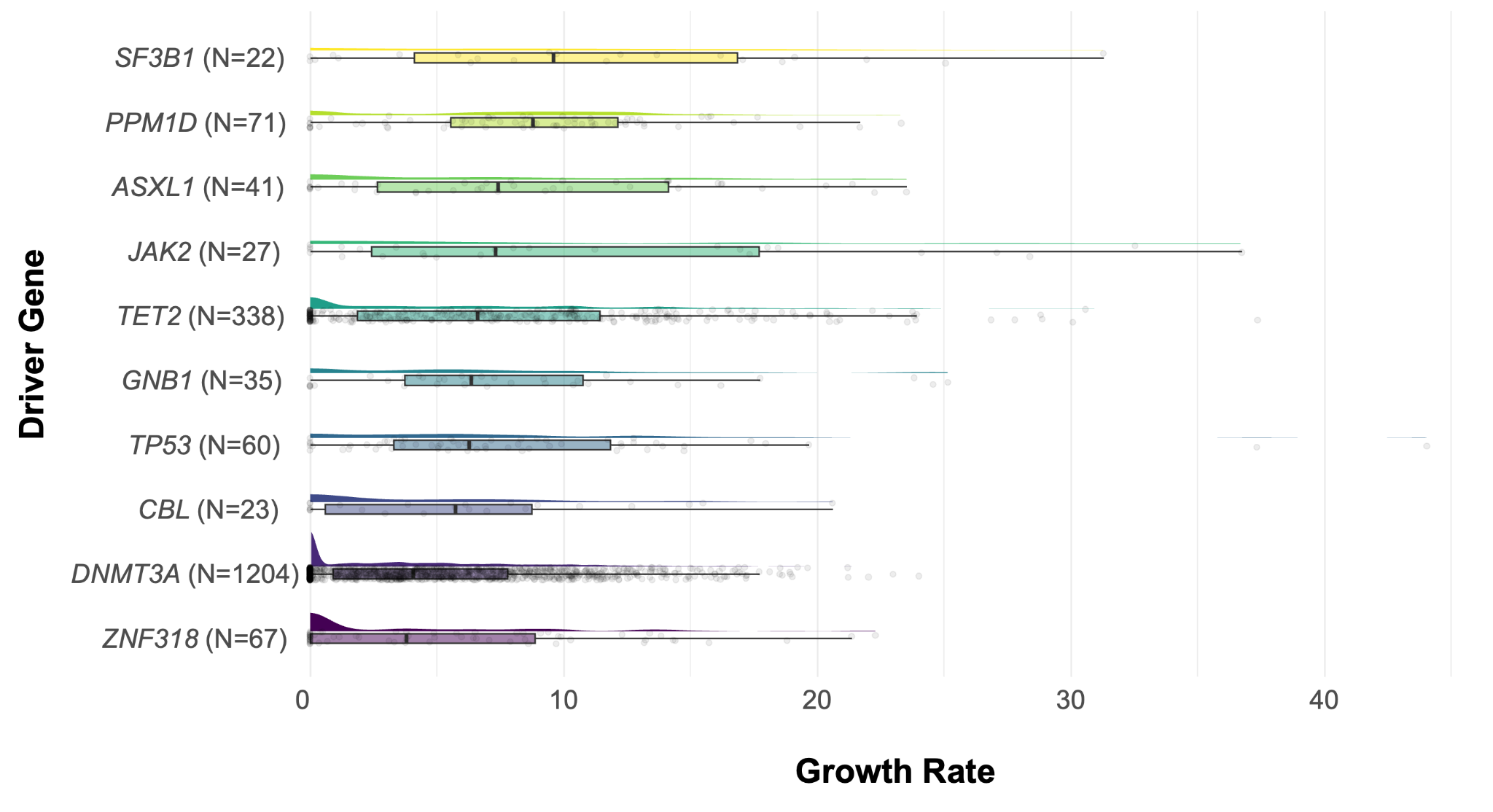

**Supplementary Figure 2:** Growth rates (%/year) of all CH mutations (VAF > 0.5%) by driver gene. Raincloud plots show distribution, with N indicating clones per gene. N represents the number of mutations with that driver gene.

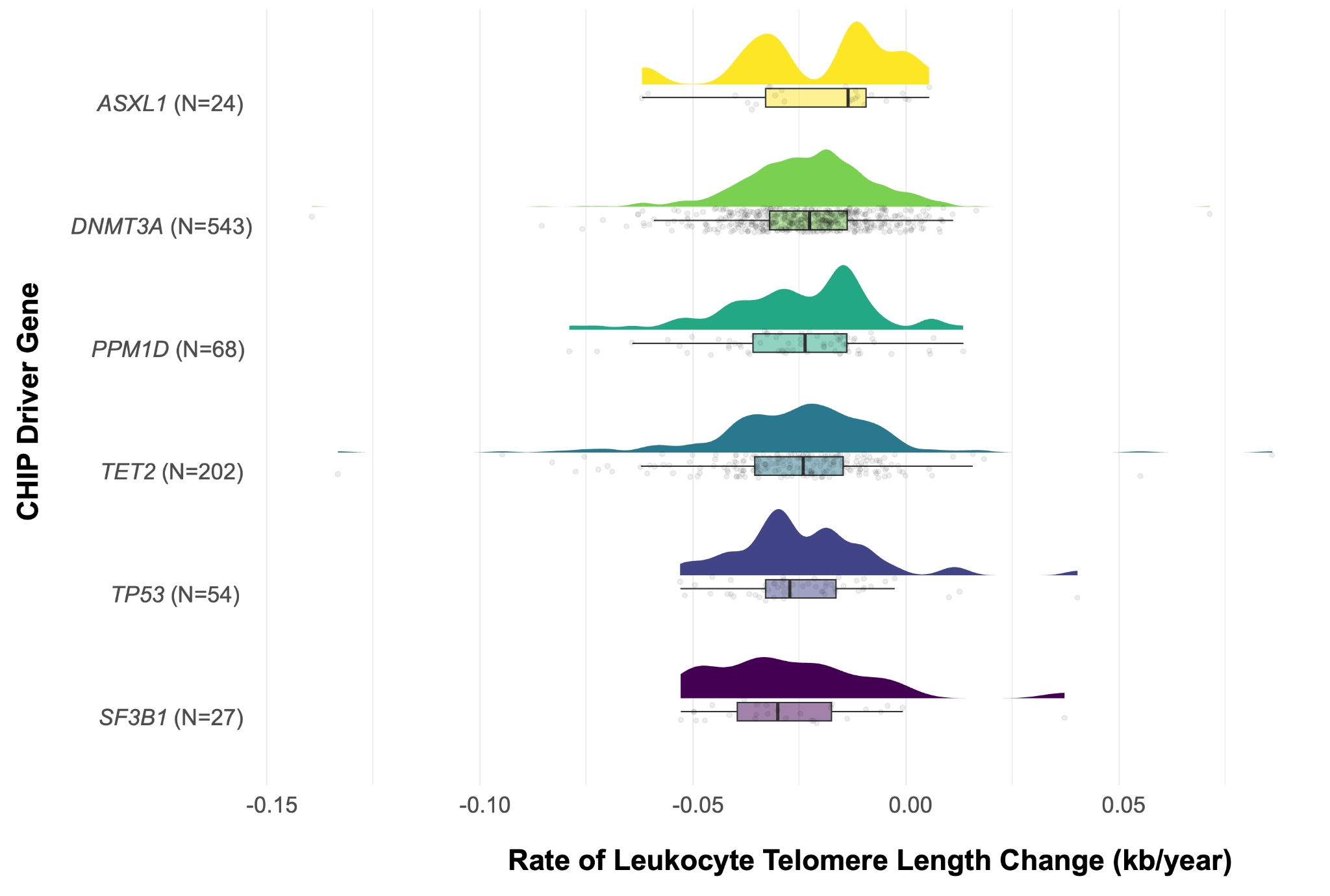

**Supplementary Figure 3:** Rate of leukocyte telomere length change (kb/year) by driver gene among participants with CHIP (VAF ≥ 2%) and leukocyte telomere length measurements at baseline and LLS. Raincloud plots show distribution, with N indicating clones per gene. CHIP: clonal hematopoiesis of indeterminate potential; LLS: Long Life Study.

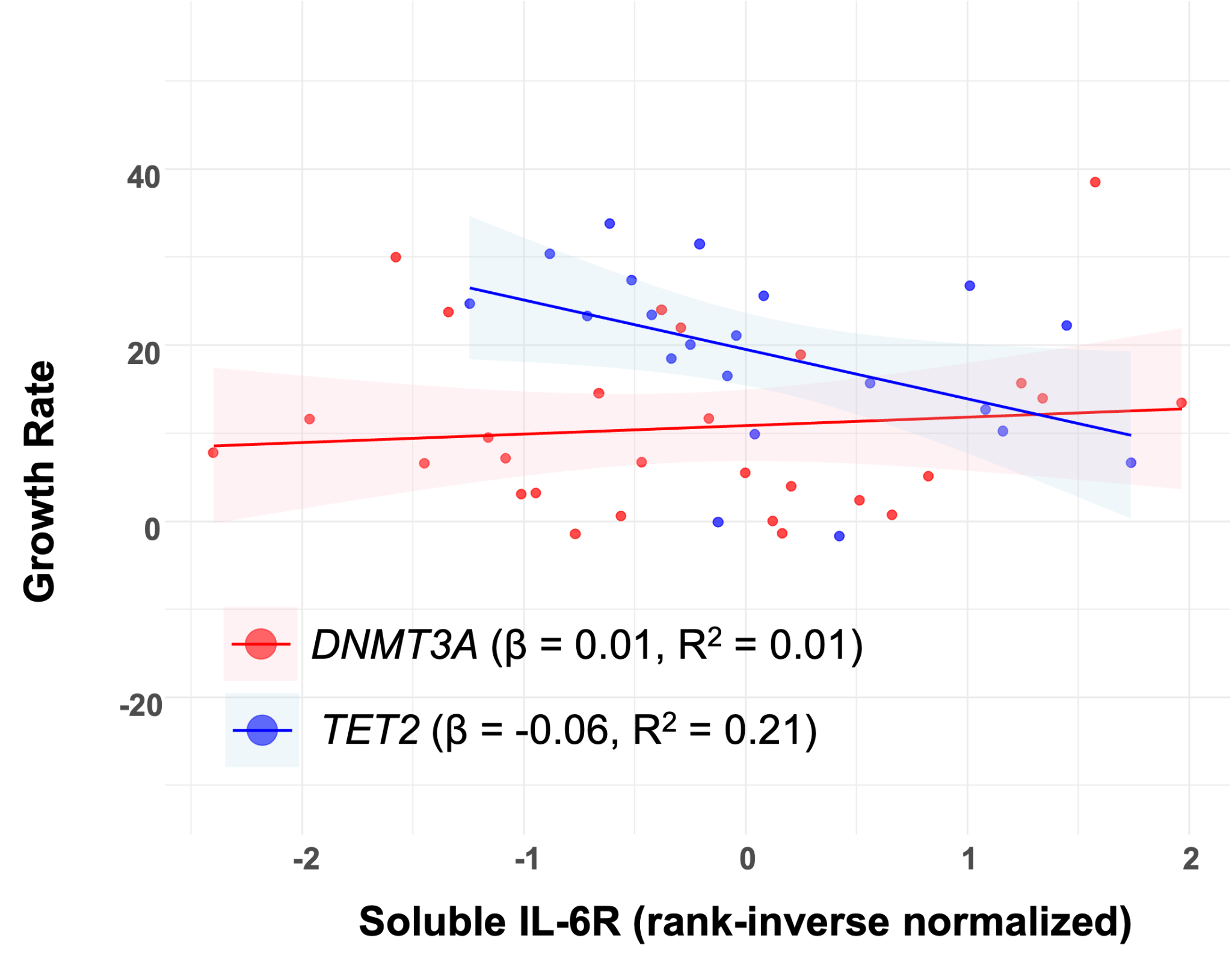

**Supplementary Figure 4: IL-6 signaling associated with *TET2* CHIP growth rate.** Scatter plot of CHIP growth rates (VAF ≥ 2%) versus soluble IL-6R levels for *DNMT3A* and *TET2* mutations. Lines represent linear regression fits with coefficients as β and correlation coefficient R² values shown.
